# Early detection of fraudulent COVID-19 products from Twitter chatter

**DOI:** 10.1101/2022.05.09.22274776

**Authors:** Abeed Sarker, Sahithi Lakamana, Ruqi Liao, Aamir Abbas, Yuan-Chi Yang, Mohammed Ali Al-Garadi

## Abstract

Social media have served as lucrative platforms for misinformation and for promoting fraudulent products for the treatment, testing and prevention of COVID-19. This has resulted in the issuance of many warning letters by the United States Food and Drug Administration (FDA). While social media continue to serve as the primary platform for the promotion of such fraudulent products, they also present the opportunity to identify these products early by employing effective social media mining methods. In this study, we employ natural language processing and time series anomaly detection methods for automatically detecting fraudulent COVID-19 products early from Twitter. Our approach is based on the intuition that increases in the popularity of fraudulent products lead to corresponding anomalous increases in the volume of chatter regarding them. We utilized an anomaly detection method on streaming COVID-19-related Twitter data to detect potentially anomalous increases in mentions of fraudulent products. Our unsupervised approach detected 34/44 (77.3%) signals about fraudulent products earlier than the FDA letter issuance dates, and an additional 6/44 (13.6%) within a week following the corresponding FDA letters. Our proposed method is simple, effective and easy to deploy, and do not require high performance computing machinery unlike deep neural network-based methods.

## 1. INTRODUCTION

As of 7^th^ September, 2021, over 220 million confirmed COVID-19 cases have been reported globally, with over 41 million reported cases in the United States (US) alone [1]. As governments and public health agencies around the globe enacted efforts to mitigate the impact of the pandemic, one persistent problem has been the opportunistic promotion of fraudulent products claiming to treat, prevent, test and/or cure COVID-19 infections. There have been numerous reports of adverse health events caused by toxic exposures to fraudulent products that have no scientific evidence supporting their use [2], [3]. In response to the emergence of many fraudulent products, the US Food and Drug Administration (FDA) has issued warning letters [4]. These warning letters are typically issued after the products become popular and many people have already been exposed to them. Since it is not possible to advertise fraudulent products on television or via reliable news sources, social media platforms have been exploited for the mass promotion of such products. In fact, promotional content regarding such products over social networks, such as Twitter, is only a subset of the misinformation spread through these platforms, which has been referred to as an *infodemic* [5], [6]. The fraudulent products are often promoted directly via the social media accounts (*eg*., Twitter, Facebook) of the entities profiting from their sales, and, if the promotions gain traction, information about them are circulated by other social media users. Consequently, information regarding the products spread through social networks in analogous patterns as other types of misinformation, including those about COVID-19 [7]. There is thus the need to develop toxicovigilance tools that can automatically identify potentially fraudulent COVID-19 products early and generate alerts. While social networks provide fertile grounds for the proliferation of misinformation about fraudulent products, they also provide opportunities for responding to diverse challenges posed by the pandemic, and one potential utility of social media is the automated real-time surveillance of fraudulent COVID-19 products.

In this paper, we demonstrate that chatter about fraudulent products on Twitter, if curated systematically via natural language processing (NLP) and data-centric methods, can provide detectable early signals. We utilize publicly available streaming data from the Twitter COVID-19 application programming interface (API), which was specifically created by the company to aid COVID-19 related research [8]. Specifically, using Twitter data, we show that social media based surveillance can detect many fraudulent products early, relative to the FDA warning issuance dates. Our approach to detecting fraudulent products is based on a simple intuition—that products that gain popularity among Twitter users, following their successful promotion, will exhibit increases in their mentions in COVID-19 related chatter. These abrupt increases in the frequency of mentions are likely to be detectable by time series anomaly detection methods. It is also likely that products that gain *relatively* higher popularity will exhibit anomalous increases of *relatively* higher magnitudes in their mentions among all COVID-19 related Twitter chatter. We describe our methods and results in the following sections.

## 2. METHODS

### 2.1 Data collection

We collected data using the COVID-19 streaming API of Twitter [8]. This API was made available by Twitter specifically for supporting COVID-19 related research, and it does not impose throughput limitations or daily/monthly quotas. Consequently, we were able to collect all tweets that mentioned the COVID-19 related keywords and phrases (*eg*., *coronavirus, covid19* and *covid*) [8]. Streaming data was stored in real-time in a *mongodb* database hosted on the Google Cloud Platform. The collection of data was continuous with only minor down times that were necessary for system modifications or updates.

### 2.2 Product detection

The list of products and entities were manually collected from the FDA website [4]. The products included were advertised as treatments/cures, tests or preventative measures for COVID-19. We curated a comprehensive list of entity names, products, FDA letter dates, person(s) who owned the entities or the products, websites and social media profiles (if any). We curated this information for a total of 183 letters issued by the FDA. Each warning letter was manually reviewed. From these, we manually curated a set of product names and/or entity names that were potentially used for promotion over social media. If the same product was mentioned in multiple letters, we only included the first mention of the product or entity and the corresponding date, excluding the later ones. We also manually curated key words and phrases that were likely to be used to refer to the products or entities on Twitter. The full list of products and entities and their earliest letter dates are provided in Table 1.

**Table 1.**
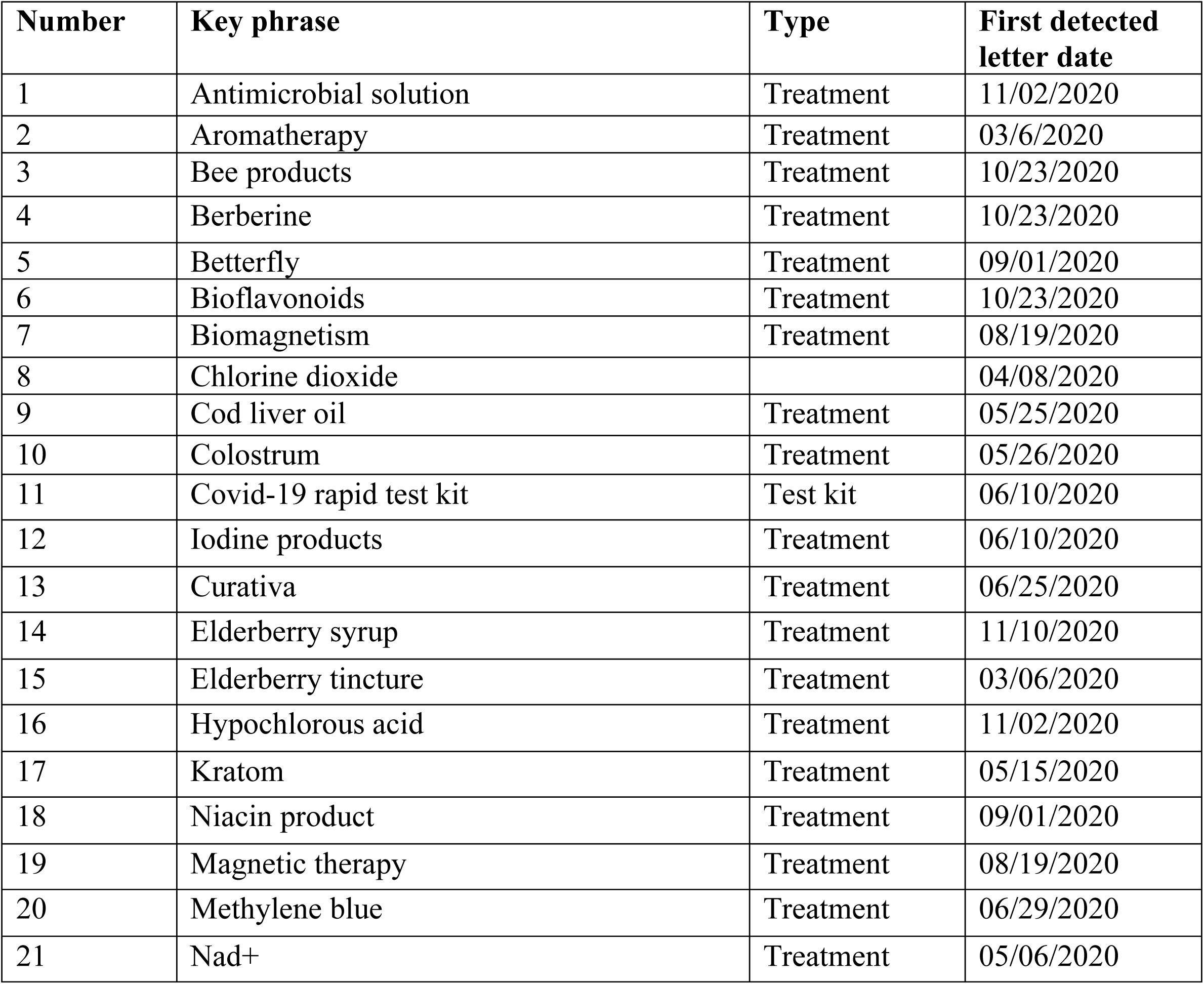
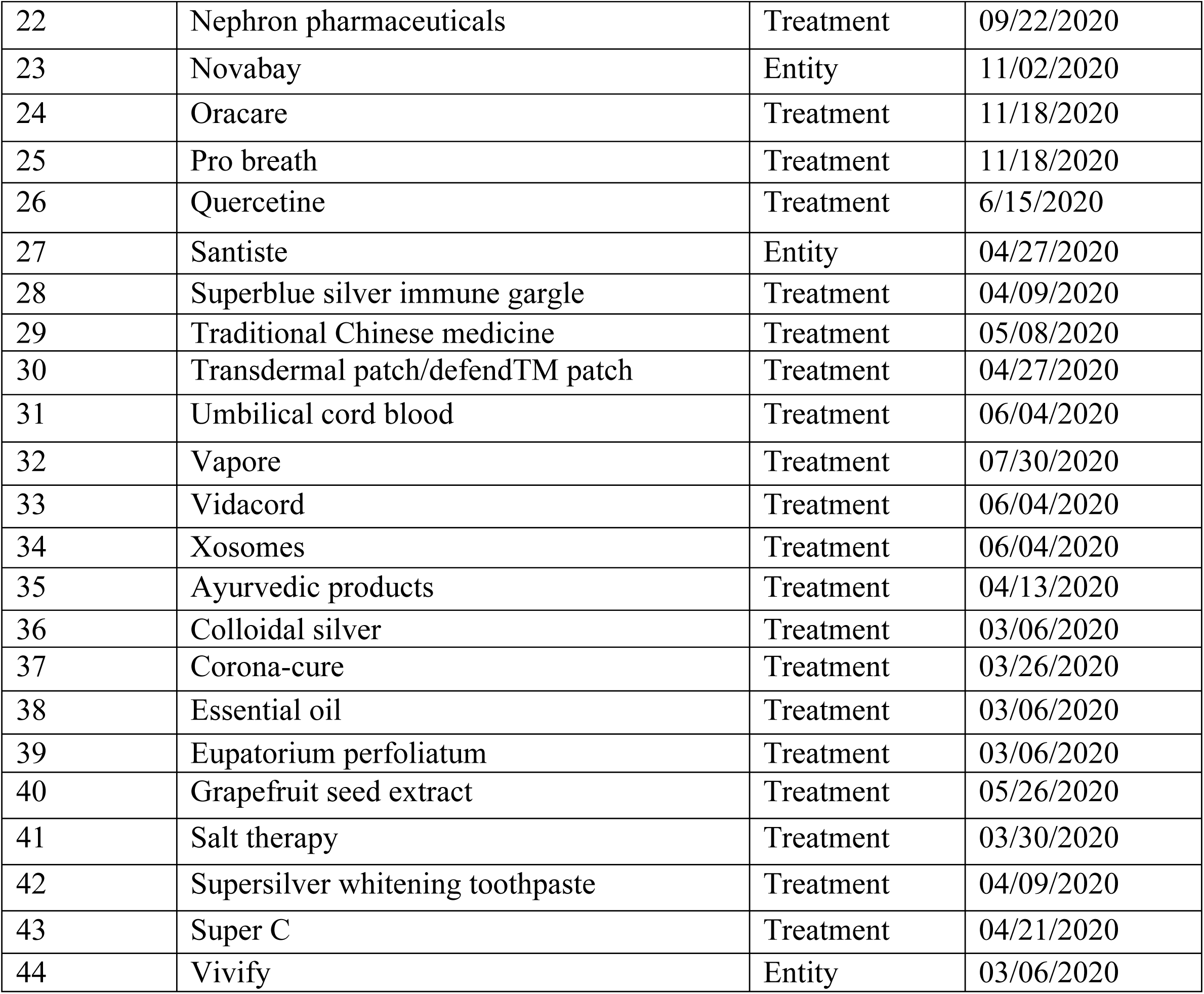
Key phrases included in this study along with their types and the date of the first letter mentioning each.

Since product and entity names are often misspelled by social media subscribers, we generated potential spelling variants or misspellings of the products and entities using a data-centric tool [9]. The variant generation tool uses a combination of semantic and lexical similarity measures to automatically identify common misspellings and spelling variants of terms/phrases, including multi-word expressions. Our past work revealed that such lexical expansion strategies are capable of significantly increasing retrieval/detection rates from Twitter [10]. Examples of product names extracted from the warning letters and their automatically-generated lexical variants are shown in Table 2. We included all products/entities and their spelling variants that had at least 10 mentions in our collected data. We excluded key phrases that were mentioned less than 10 times because such low occurrences indicated that the corresponding products/entities were either not promoted over Twitter or never actually gained popularity on the platform. We counted the number of mentions of each product/entity, including their spelling variants, from the entire collected dataset. Counts of spelling variants were grouped with the original products/entities. Daily counts were normalized by the total number of posts collected on the same days. The daily relative frequencies were represented as the number of mentions per 1000 tweets.

**Table 2.**
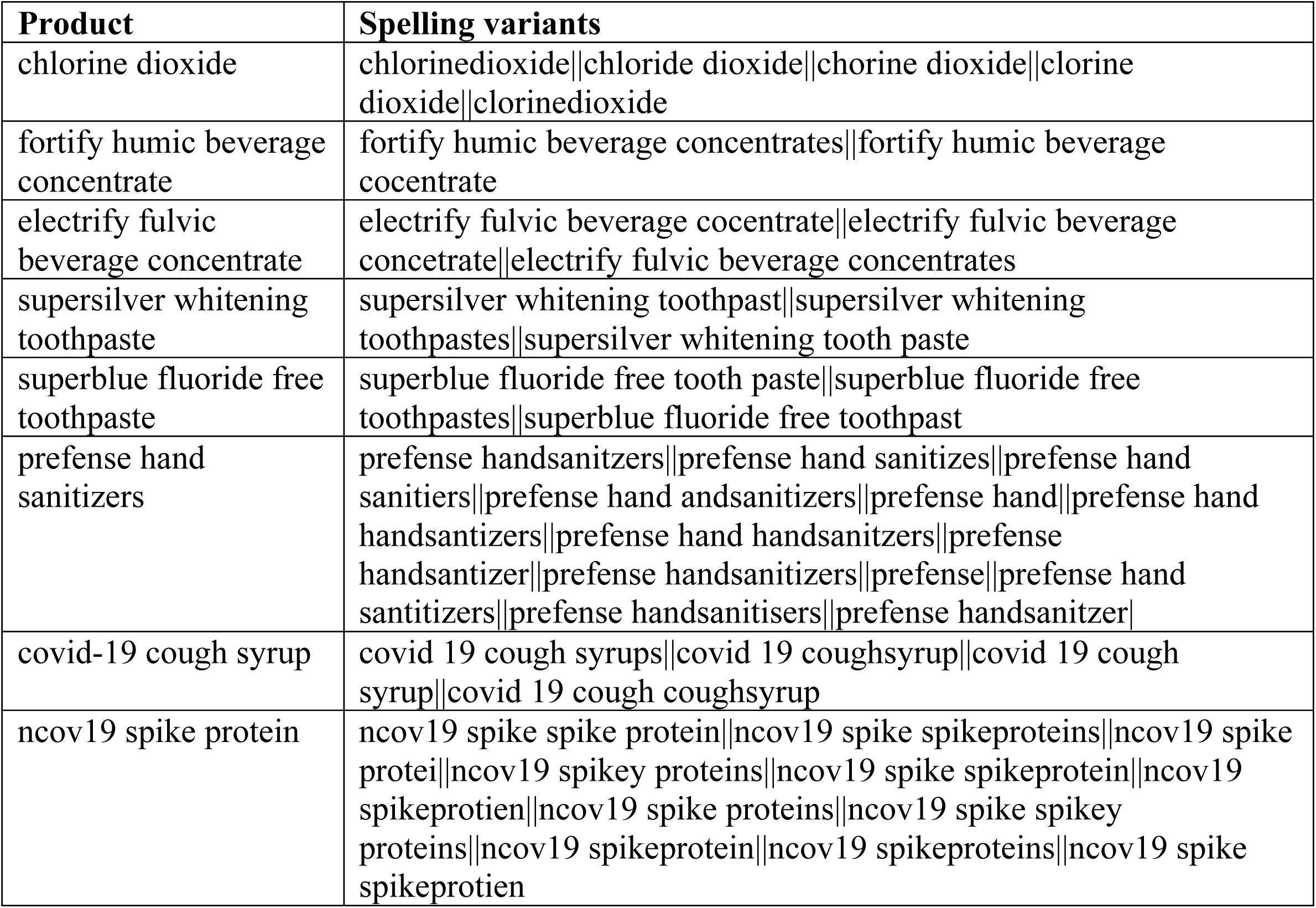
Fraudulent product names extracted from the FDA warning letters and their automatically-generated lexical variants.

### 2.3 Detecting anomalies

We applied a 14-day moving average filter to construct a smooth line representing the daily mention frequencies, and anomalies or outliers were detected relative to this moving average line. For each day, the *residual* for standard deviation calculation was computed by subtracting the 14-day moving average from the relative frequency per 1000 tweets on that day. For a given day (***n***), the standard deviation for the day (***σ***_***n***_), is computed progressively, given as:

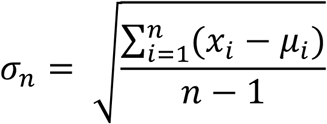

where ***x***_***i***_ is the relative frequency for day ***i*** and ***µ***_***i***_ is the 14-day moving average on day ***i***. Thus, the standard deviation computed for a given day includes all the data points starting from day 1. The standard deviation for the first day (February 19, 2020) for any product is by definition 0. This may potentially give the anomaly detection approach an unfair advantage by increasing the sensitivity of detection in the early days easier. Therefore, we artificially added a non-zero standard deviation on day 1, computed as ^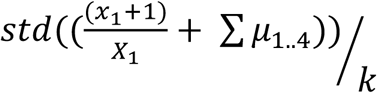^, where ***x***_**1**_is the product mention frequency on day 1, ***X***_**1**_is the total number of tweets collected on day 1, ***std*()** is the standard deviation function, and ***µ***_**1**..**4**_, are the moving averages over the first 4 days. The value is divided by ***k*** to adjust for the ***k* × *std*(**) function that is applied to compute the boundaries beyond which a data point would be considered an outlier (***k***=3 in our experiments). This artificial initial bias that we added therefore decreases the chances of our approach to detect outliers early on in the timelines, and makes the task of detecting anomalies slightly harder, particularly for products that have relatively low number of mentions. For some products, for example, there are many days with 0 mentions early on in their timelines, but the added bias causes the progressive standard deviation to be non-zero. For 3 products with letter issue dates in March 2020, this added bias caused the method to miss early outliers that are detectable without adding the bias. Specific details are provided in the supplementary material (S3). Minimum value for daily relative frequency was set at 0.001 (*i.e*., ***k* ×** 0.001 served as the minimum threshold for outlier detection).

The chosen window size (14) and standard deviation (3), for which we report results in this paper, were relatively conservative choices for signal detection. We also performed experiments with multiple window sizes (7, 10, and 14) and standard deviation thresholds (2, 2.5, and 3) to study how the anomaly detection performance varied based on these parameters. Slight variations in window sizes and standard deviations did not impact overall performance.

### 2.4 Evaluation

Data points that had a distance of more than 3 standard deviations from the moving average were considered to be outliers (*i.e*., signals). For each key phrase, the date of the first outlier was compared with the FDA letter issuance date to determine if the signal was detected earlier, within 1 week, or later than the FDA letter issuance date. System percentage accuracy was computed using the formula: *#early/#total*. For products that were mentioned in multiple letters, our approach was only considered to be successful in early detection if the outlier was detected prior to the first mention date. Thus, the reported system performance is actually likely to be lower than in practice.

## 3. RESULTS

The issue dates of the letters ranged from March 6, 2020 to June 22, 2021. Through manual review of each letter, we identified 221 potential keywords/phrases that were either associated with the products (*e.g*., product names) or the entities selling them. From this set, we excluded key phrases collected after the year 2020. Some products were promoted by different entities at different times, causing them to be repeated in the warning letters. Since our primary objective was to assess the possibility of early detection, we excluded repeated key phrases, keeping only their first occurrences (n=56). Furthermore, since our focus was to detect products that gained popularity via promotion on Twitter, we excluded key phrases that were mentioned less than 10 times including their lexical variants (n=12). 44 key phrases met all the inclusion criteria. Table 1 presents all 44 keywords, their types (*ie*., product or entity), and the FDA letter issuance dates. The full curated data along with additional information is available as supplementary material (Table S1).

We included a total of 577,872,350 COVID-19 related tweets in our analysis, which were collected from February 19, 2020 to December 31, 2020. We computed the daily counts of the key phrases (along with their spelling variants, if any). Increases in key phrase mention counts that were higher than 3 standard deviations from the 14-day moving average of mentions were flagged as potential ‘signals’. 43 out of the 44 key phrases showed anomalous increases in their mentions at some point of time within our collected data. For 34 out of the 44 key phrases (77.3%), signals of anomalous increases in chatter were detectable prior to the FDA letter issuance dates. An additional 6 (13.6%) key phrases had anomalous increases within seven days of the FDA letter issuance dates. Daily counts for the products, their 3-standard deviation ranges, and the moving averages are shown in Figure 1 of supplementary material. The figure specifically shows the daily normalized frequencies for each product from 02/19/2020 to 12/31/2020. 14-day moving average is represented by a blue line and the range of three standard deviations is shown as a shaded area. A vertical line in each figure (red) shows the FDA letter issuance date and orange-black circles represent the data points falling outside the three standard deviation range. Products detected earlier than the FDA letter issuance dates are indicated by a lime-green background and the rest are indicated by a beige background. The ordering of the products is the same as the ordering in Table 1. The daily counts for all 44 key phrases are provided as supplementary material (S2; daily**_**counts.xlsx).

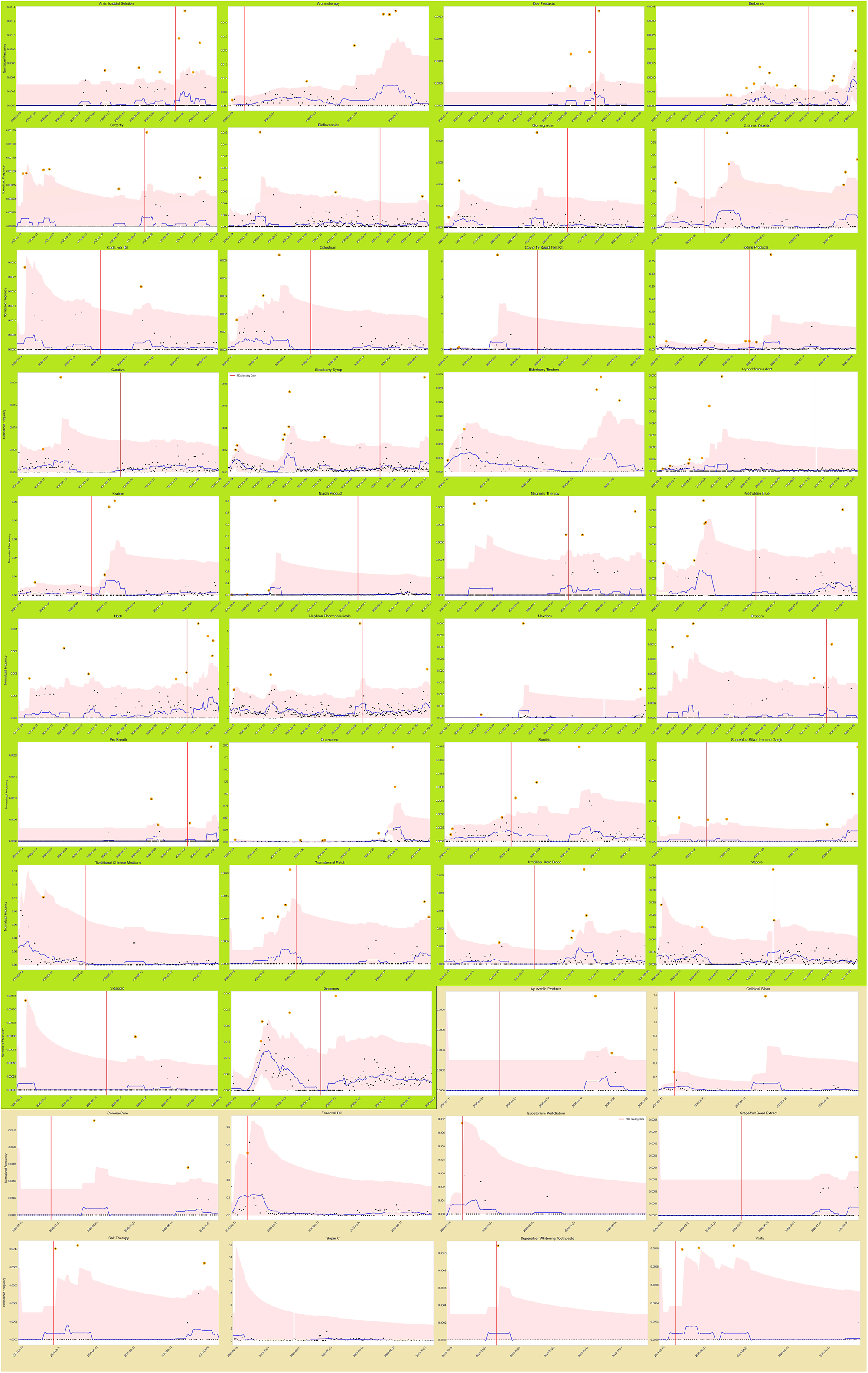

## 4. DISCUSSION

### 4.1 Related work

Our work is not the first to explore the utility of social media as a potential source for detecting fraudulent COVID-19 products. In recent works, unsupervised NLP methods such as topic modeling and supervised methods such as text classification have been proposed for the automatic detection of such products from social media data [11]–[13]. Others focused more broadly on detecting misinformation using social media or Internet-based data [14], [15]. However, these studies did not take into account the time factor. Typically, once the FDA issues warning about a fraudulent product, there is a rise in chatter regarding the product, but such rises are driven by media coverage or increased public awareness. We observed this phenomenon for most products included in the study, particularly the ones detected within one week of the FDA letter issuance dates. To the best of our knowledge, our approach is the first to attempt to detect fraudulent treatments early. The proposed approach is also simple, computationally inexpensive as it relies on fundamental characteristics of social media chatter (*ie*., increases in volume of chatter about a particular topic resulting from increases in its popularity), and unsupervised (*ie*., no training data required).

### 4.2 Limitations

There are several potential limitations of the proposed approach. First, it requires data that is not rate limited (*eg*., data from the standard Twitter streaming API). Anomalous increases may not be detectable from rate limited streams since large increases in volume are likely to be dampened by the APIs. For real-time fraudulent product candidate detection, deployment needs to be on streaming data, although it is also possible to periodically run the anomaly detection scripts on stored, static data. Second, we were only able to calculate the percentage of early detection within our given sample, and based on the current data, we were unable to realistically estimate confidence intervals for the percentage values reported. Third, the anomaly detection approach relies on characteristic abrupt increases in chatter volumes about a given topic. It is possible that some fraudulent products may gain popularity gradually, causing the normalized counts to never go beyond the standard deviation threshold. In such cases, varying the window size (*eg*., using 7-day moving averages) and/or lowering the standard deviation thresholds may improve the detection capability of the method. However, lowering the standard deviation threshold is also likely to result in larger numbers of false positives—an aspect that we did not take into account in this study. We believe that not taking false positives into account in the current study is justifiable since in practical settings, all signals associated with noun phrases would be reviewed by experts, and so it is perhaps better if the method is biased in favor of *recall* (*ie*., more true and false positives) rather than *precision*.

We also do not address candidate fraudulent substance detection in this study. Several mechanisms can be used for detecting candidates including but not limited to named entity recognition (likely to be high precision but low recall), simple part-of-speech tagging to identify noun phrases (high recall, low precision), and topic modeling methods that identify possible topics from texts (low recall, high precision). We intend to explore these strategies in future work. Even without this component, we believe our approach is an improvement over past studies that did not take into account the warning letter dates. Finally, since the daily counts are normalized by the total number of tweets on the same day, it is possible that large increases in absolute counts of specific key phrases are not detectable due to equal or larger increases in the total volume of posts on the same day.

## 5. CONCLUSION

The emergence of fraudulent products associated with COVID-19 has been a significant problem in the fight against the pandemic. Social media has served as platforms for advertising and promoting fraudulent products. While social media makes it easier for opportunist entities to promote and sell fraudulent products, this resource may also be used to conduct surveillance of fraudulent substances. In this paper, we showed that it is possible to detect many fraudulent products potentially early from Twitter data. Our simple approach employed a time series anomaly detection method for detecting anomalous increases in mentions of fraudulent substances in Twitter chatter, and obtained promising performance. Future work will focus on deploying the NLP pipeline and improve upon the limitations described in the Discussion section.

## Data Availability

Available as supplement

## CONTRIBUTIONS

AS conceived the study, designed the experiments and prepared the manuscript. SL conducted data collection, experiments and image generation. RL and AA prepared and reviewed the dataset. YY and MA to review the research and help with preparation of the manuscript.

