## Supplementary material for "Early detection of fraudulent COVID-19 products from Twitter chatter": Supplmentary material.docx

**Table S1.** Full curated details from FDA warning letters.

| **Letter date** | **Company** | **Product** | **Person** | **Website** |
| --- | --- | --- | --- | --- |
| 3/6/20 | Xephyr LLC dba N-ergetics | Colloidal silver products, including Colloidal Silver 1100 PPM | Brad Brand, Derill J. Fussell, and Linda Fussell |  |
| 3/6/20 | GuruNanda, LLC | Essential oil products | Mr. Puneet Nanda | www.gurunanda.com |
| 3/6/20 | Quinessence Aromatherapy Ltd | Essential oil products | Geoff Lyth | www.quinessence.com |
| 3/6/20 | Vivify Holistic Clinic | Formula #1, Formula #2, Formula #3, and Eupatorium perfoliatum (Boneset) | David Raes | https://coronavirusdefense.com |
| 3/6/20 | Colloidal Vitality LLC | Products labeled to contain silver | Jennifer Hickman | www.purevitalsilver.com |
| 3/6/20 | The Jim Bakker Show | Silver Sol Liquid and products labeled to contain silver | James Bakker | www.jimbakkershow.com |
| 3/6/20 | Herbal Amy Inc. | Coronavirus Protocol (Coronavirus Boneset Tea, Coronavirus Cell Protection, Coronavirus Core tincture, Coronavirus Immune System, and Elderberry Tincture) | Amy Weidner | www.herbalamy.com |
| 3/26/20 | Corona-cure.com | Corona-Cure Coronavirus Infection Prevention Nasal Spray |  | https://corona-cure.com |
| 3/26/20 | Carahealth | Herbal products, including "Carahealth Immune" also referred to as "Immune Tonic" | Carina Harkin | www.carahealth.com |
| 3/30/20 | Halosense Inc. | Salt therapy products | Livia Tiba,Director/CEO | www.salinetherapy.com |
| 3/30/20 | JRB Enterprise Group Inc. DBA Anti Aging Bed | Colloidal silver products | Mr. John Baxter | www.antiagingbed.com |
| 3/30/20 | Bioactive C60/FullerLifeC60 LLC | "FullerLifeC60" | Marshall Thurber | www.bioactivec60.com |
| 3/31/20 | Neuro XPF | Cannabidiol (CBD) products |  | https://neuroxpf.com |
| 4/1/20 | Homeomart Indibuy | Homeopathic drugs | Mr. Vasant Prabhu | www.homeomart.com |
| 4/1/20 | Gaia's Whole Healing Essentials, LLC | Colloidal silver products | Mr. Michael Lamboy | www.gaiaswholehealingessentials.org |
| 4/1/20 | Health Mastery Systems DBA Pure Plant Essentials | Essential oil products | Ms. KG Stiles | www.pureplantessentials.com |
| 4/6/20 | Ananda, LLC DBA Ananda Apothecary | Essential oil products | Mr. Scott Merrill | www.anandaessentialoils.com |
| 4/6/20 | Alternative Health Experts LLC DBA Immunization Alternatives | Homeopathic drugs and dietary supplement products |  | www.homeopathicremediesonline.com |
| 4/6/20 | Cathay Natural, LLC | Herbal products including "CoronaDefender Herbal Sachet-S" | Ms. Cathay Fung | www.cathayherbs.com |
| 4/6/20 | Native Roots Hemp | Cannabidiol (CBD) products |  | https://farm2pharmacy.com,https://www.facebook.com/NativeRootsHemp/, |
| 4/6/20 | Indigo Naturals | Cannabidiol (CBD) products |  | https://indigonaturals.net, www.facebook.com/IndigoNaturals/, |
| 4/7/20 | CBD Online Store | Cannabidiol (CBD) products |  | https://cbd-online-store.com/ |
| 4/7/20 | Savvy Holistic Health dba Holistic Healthy Pet | "China Oral Nosode,” also called “China Corona Nosode," and the “CV Respiratory Kit,” under the description “AN330 – CORONA VIRAL IMMUNE SUPPORT AND/OR ACTIVE RESPIRATORY INFECTION FOR ALL AGES” |  | https://holistichealthypet.com/ |
| 4/8/20 | Genesis 2 Church | Miracle Mineral Solution (MMS) (chlorine dioxide) | Mark Grenon, Joseph Grenon, Jordan Grenon, and Jim Humble | https://genesis2church.is, https://genesis2church.ch, https://g2churchnews.org, https://g2voice.is, and https://jimhumble.co |
| 4/8/20 | NRP Organics Ltd | "Fortify Humic Beverage Concentrate” and “Electrify Fulvic Beverage Concentrate” | James Rutherford | www.coronaviruscures.com |
| 4/9/20 | Free Speech Systems LLC d.b.a. Infowars.com | “Superblue Silver Immune Gargle,” “SuperSilver Whitening Toothpaste,” “SuperSilver Wound Dressing Gel” and “Superblue Fluoride Free Toothpaste” | Mr. Alexander E. Jones | www.infowarsstore.com |
| 4/9/20 | Earthley Wellness dba Modern Alternative Mama LLC | Herbal tinctures and herbal remedy products |  | https://earthley.com and https://modernalternativemama.com |
| 4/10/20 | Herbs of Kedem | Herbal products |  | http://kedemnatural.com |
| 4/13/20 | Gaia Arise Farms Apothecary | “True Viral Defense,” also referred to as “Viral Defense Tincture” | Nicole Mescia | https://gaiaarisefarmsapothecary.com |
| 4/13/20 | The GBS dba Alpha Arogya India Pvt Ltd | Ayurvedic products (“Alpha 11” and “Alpha 21”) | Ajay Goyal,CEO/Founder | www.alphaarogya.com |
| 4/14/20 | Earth Angel Oils | Essential oil products | Helen Stembridge | www.earthangeloils.com |
| 4/15/20 | The Art Of Cure | Homeopathic drug products |  | https://theartofcure.net/ |
| 4/16/20 | Nova Botanix LTD DBA CanaBD | CBD products |  | https://canabd.com/ |
| 4/21/20 | Copper Touch, LLC | "Sani-Bar GK95” and “Sani-Disc GK95D” | Linda Pavek | https://coppertouch.com |
| 4/21/20 | DrJockers.com, LLC | Super C, Vitamin D/K2, Zinc Charge, and ImmunoStrong Berry Liquid | David Jockers | www.drjockers.com |
| 4/23/20 | Prefense LLC | Prefense Hand Sanitizers | Mr. Michael Reiber | www.prefense.com |
| 4/27/20 | Santiste Labs LLC | “DefendTM Patch” |  | https://defendpatch.com |
| 4/27/20 | Hopewell Essential Oils | Essential oils and herbal products |  | https://hopewelloils.com |
| 5/4/20 | Honey Colony LLC | "Quicksilver Liposomal Vitamin C w/ Liposomal,” “Jigsaw Magnesium With SRT,” and products labeled to contain silver, including “Silver Excelsior Serum” | Maryam Henein | www.honeycolony.com |
| 5/4/20 | Dr. Dhole's Sushanti Homeopathy Clinic | “Homeopathic Genus Epidemicus” | Dr. Nitin S. Dhole | www.homeopathyhelps.com |
| 5/6/20 | GlutaGenic | Viral Protection Kits |  | www.glutagenic.com and www.advancedbiomecorp.com |
| 5/6/20 | Alive By Nature, Inc. | “NAD+” and “NMN” sublingual gel products |  | https://alivebynature.com/,www.facebook.com/alivebynature/, |
| 5/7/20 | Sanit Technologies LLC dba Durisan | Non-alcohol based hand sanitizer products | Mr. Troy M. Daland,CEO | https://durisan.com |
| 5/7/20 | AgroTerra, Ltd. dba Patriot Hemp Company | CBD products, silver, iodine, medicinal mushroom, vitamin C, selenium, zinc, vitamin D3, astragalus, and elderberry | Mr. Doug Pineda | www.patriotcbd.org |
| 5/7/20 | WashingtonsLastFrontier.Com | Essential oils and dietary supplements | Ms. Tiffany Davidson | www.washingtonslastfrontier.com |
| 5/7/20 | Chronic Lyme Treatments | Herbal products, including "COVID-19 Core Formula," "Immunity Blend," and "Protection Blend" | Sal Meli | www.chroniclymetreatments.com |
| 5/8/20 | Seanjari Preeti Womb Healing, L.L.C. | Honey product, "COVID-19 COUGH Syrup" | Ivy Ruffin (Sagarius) | www.seanjaripreeti.com |
| 5/8/20 | Plum Dragon Herbs, Inc. | Traditional Chinese medicine (TCM) products | Lisa Ball | www.plumdragonherbs.com |
| 5/11/20 | Fusion Health and Vitality LLC | "CORE” and “IMMUNE SHOT” | Polly Ryncarz | www.pharmorigins.com, http://www.freecoreoffer.online, and http://www.immune-shot.com |
| 5/11/20 | Center for New Medicine/Perfectly Healthy by Connealy MD | Camu Camu’s Vitamin C, Vitality C, Immunity Kit, Vitamin D3-K2, Liquid Silver, and Power Immune | Leigh Erin Connealy, M.D. | www.perfectlyhealthy.com, www.cfnmedicine.com, www.cancercenterforhealing.com, and www.shopcfnmedicine.com |
| 5/14/20 | White Eagle Native Herbs | Herbal products | Mark White Eagle | https://www.whiteeaglenativeherbs.net |
| 5/14/20 | benjaminmcevoy.com | Dietary supplements | Benjamin McEvoy | www.benjaminmcevoy.com |
| 5/15/20 | Noetic Nutraceuticals | CBD products |  | www.nnlifestyle.com |
| 5/15/20 | The Golden Road Kratom | Kratom products |  | www.thegoldenroadkratom.com |
| 5/15/20 | Natural Solutions Foundation | "Dr. Rima RecommendsTM Nano Silver 10 PPM” products |  | www.drrimatruthreports.com, www.opensourcetruth.com, www.truthaboutcoronavirus.com, and www.nsfmarketplace.com |
| 5/19/20 | SpiceTac | Vitamin C product |  | www.spicetac.com |
| 5/20/20 | Life Unlearned, LLC | Vitamin D products |  | www.lifeunlearned.com |
| 5/21/20 | Apollo Holding LLC | “NoronaPak” products, including cannabidiol (CBD) and other supplement products | Bryce Johnson | https://apollohempire.com and https://www.noronapak.com |
| 5/21/20 | North Coast Biologics | “nCoV19 spike protein vaccine” | Johnny T. Stine | www.northcoastbio.com |
| 5/26/20 | Careful Cents, LLC | Essential oil products | Carrie Lee Smith Nicholson | https://abeginnersguidetoessentialoils.com |
| 5/26/20 | Musthavemom.com | Colloidal silver, vitamins, minerals, herb oils and a homeopathic drug product |  | www.musthavemom.com |
| 5/26/20 | Alternavita | Grapefruit seed extract, colostrum, and cod liver oil products | Mary Ferrari | www.alternavita.com |
| 5/26/20 | CBD Gaze | CBD products including Restorative Botanicals Ultra High Strength Hemp Oil Supplement and Serenity Hemp Liquid Dietary Supplement |  | www.cbdgaze.com |
| 5/28/20 | Quadrant Sales & Marketing, Inc. | Non-alcohol based hand sanitizer product | Robert Cefail,President | https://cleanhandsandmore.com/ |
| 5/28/20 | StayWell Copper Products | “Germ Stopper” products | Marcia Reece & Diana French | www.staywellcopper.com |
| 6/1/20 | Dr. Sherrill Sellman | HealthMax Nano-Silver Liquid, Silver Biotics Silver Lozenges with Vitamin C, and Silver Biotics Silver Gel Ultimate Skin & Body Care (collectively, “your silver products”) | Dr. Sherrill Sellman | www.drsherrillsellman.com, archive.aweber.com/whatwomenmust, and www.whatwomenmustknow.podbean.com |
| 6/4/20 | EUCYT Laboratories LLC | Products derived from human umbilical cord blood and umbilical cord, VidaCord™, VidaGel™ and VidaStem™; an exosome product, XOsomes™; and an amniotic fluid derived product, VidaFlo™ | Travis H. Bird,Chief Executive Officer | https://eucyt.com |
| 6/8/20 | organic-beauty-recipes.com | Essential oil products | Eve Cabanel | www.organic-beauty-recipes.com |
| 6/10/20 | www.outoftheboxremedies.com | Iodine products | John Burket | www.outoftheboxremedies.com, www.outsidetheboxremedies.yolasite.com, and www.outoftheboxremedies.net |
| 6/10/20 | Medakit Ltd | "COVID-19 Rapid Test Kit” (aka “COVID-19 IgM/IgG Rapid Test” and “Covid-19 Fast Test Kit”) |  | www.medakit.com |
| 6/15/20 | antibodiescheck.com | “Antibodies Test Kit for Covid19” (aka “Corona Check Kit,” “Antibodies Check Kit,” “Covid-19 IgG/IgM Rapid Test Cassette (WB/S/P),” and “Corona Antibodies Test” |  | www.antibodiescheck.com |
| 6/15/20 | Sonrisa Family Dental dba www.mycovidtest19.com | “Cellex Test Kit” and “Leccurate Test Kit” | Dr. Jason Korkus, DDS | www.mycovidtest19.com |
| 6/15/20 | Sovereign Laboratories, LLC | “PRO Vital C-LD®,” “PRO Colostrum-LD®,” “Vital C-LDTM,” and “Colostrum-LD®” | Douglas Wyatt | www.sovereignlaboratories.com |
| 6/15/20 | FRS International, LLC | Quercetin-containing soft chew and liquid concentrate products | Thomas C. Lines | www.frs.com, www.healthyenergy.com and www.frs-international.com |
| 6/17/20 | KBMO Diagnostics, LLC | “COVID-19 Fingerstick Test Kit” | James White,CEO | www.kbmodiagnostics.com |
| 6/17/20 | Modern Allergy Management LLC dba Direct Med Solutions LLC | "COVID-19 Fingerstick Test Kit" | George Masey | https://covid.directmedsolutions.com/fingerstick |
| 6/18/20 | Project 1600 Inc. | Cannabidiol (CBD) products | Dennis Michael Lynch | https://dmlcbd.com |
| 6/19/20 | North Isle Wellness Center | Methylene Blue products, including Blu Block |  | https://theblublock.com/ |
| 6/25/20 | Curativa Bay Corporation | Advanced Hypochlorous Skin Spray | William J. Maher | https://www.curativabay.com |
| 6/26/20 | Nuance Health, LLC | “Swype Shield” | Kent New and Jennifer Sawyer New | https://swypeshield.com and www.swype-shield.myshopify.com |
| 6/26/20 | SuperHealthGuard and Loyal Great International Ltd. | Lianhua Qingwen Capsule |  | www.superhealthguard.com |
| 6/29/20 | Epro E-Commerce Limited dba DealExtreme and DX.com | “Wondfo Novel Coronavirus (2019-nCoV) Antibody Detection Kit” | Xiao Fei Che | https://www.dx.com |
| 6/30/20 | Center for Wellness and Integrative Medicine | "COVID Supplement Protection Pack” (also referred to as the “COVID Household Value Pack”), Thymosin-Alpha, and Methylene Blue Capsules | Tom Yarema, M.D. | https://drtomyarema.com |
| 7/6/20 | Shen Clinic, LLC | Traditional Chinese Medicine (TCM) products, including "SHUANG HUANG LIAN" and "Gan Mao Ling" |  | https://shenclinic.com |
| 7/6/20 | Lianhuaqingwencaps.com | Traditional Chinese medicine (TCM) product, “Lianhua Qingwen” |  | https://lianhuaqingwencaps.com/ |
| 7/6/20 | Lotus Herbal Supplements | Traditional Chinese medicine (TCM) product, “Lianhua Qingwen Capsules” |  | www.lotusherbalsupplements.com |
| 7/6/20 | Butterfly Expressions LLC | Blessed waters, essential oils, hand sanitizers, homeopathic products, and tinctures | Laree Westover & Valaree Westover (Sharp) | www.butterflyexpress.shop |
| 7/6/20 | SinoTradition.com | Traditional Chinese medicine (TCM) products, including "Lianhua Qingwen Capsules" and "Qing Fei Pai Du Tang" |  | www.sinotradition.com |
| 7/7/20 | Ionogen, LLC | “Ionopure Skin & Hands” |  | ionopure.com |
| 7/10/20 | Health Beauty Love | Tincture, “E-Munity" |  | https://healthbeautylove.com/ |
| 7/13/20 | Kegan Wellness | “She Vitamin C Tablets,” “She+ Tablets,” and “Giloe+ Tablets” |  | https://www.keganwellness.com |
| 7/21/20 | 21st Century LaserMed Pain Institute d/b/a Create Wellness Clinics | Umbilical cord derived stem cell product, Exosome product, Deluxe Immune Support Bundle” (Immune Support Bundle), “Dr. Phillip Yoo D.C.’s Signature – Emergency II COR-1:9 CV-19 Deluxe Immune Boost Protection Kit,” and “Create Wellness Clinics COVID-19 Coronavirus SARS-CoV-2 Antibody At Home Rapid Telemedicine10 Minute Telemedicine Test Kit With Virtual Consultation” | Dr. Phillip Yoo, DC | www.laserstemed.com, www.lasermedpaininstitute.com, www.laserstemproducts.com, and www.drphillipyoo.com |
| 7/23/20 | AkivaMed Inc. | “COVID-19 Antibody Rapid Test Kit” |  | www.akivamed.com |
| 7/23/20 | Holistic Health International, LLC | At-home stool sample collection kit, “SARS-COV-2 STOOL TEST (SCV)” | Dr. Amy Yasko and Edward Yasko | www.holisticheal.com and https://www.ch3nutrigenomics.com/ |
| 7/24/20 | CoreMedica Laboratories, Inc. | At-home sample collection kit, “COVID-19 (SARS-CoV-2) ANTIBODY SCREENING KIT Specimen Self-Collection & Transport Kit” | Cory Zuehlsdorf,COO & Director North America | www.coremedicalabs.com and www.testmydrop.com |
| 7/24/20 | Fair Price Labs, Inc. | At-home sample collection kit | Deryl Everette and Patricia Everette | www.fairpricemd.com |
| 7/30/20 | Vapore LLC dba Mypurmist | Mypurmist |  | https://www.mypurmist.com/ |
| 8/3/20 | MMSTabs.com | MMS Products | Terance Winson | www.mmstabs.com |
| 8/6/20 | Canadian Chaga | "Chaga products" including "124 Chaga Capsules,” “Chaga Tea,” and “Canadian Chaga Tincture” |  | https://canadianchaga.com,https://canadianchaga.com/ |
| 8/7/20 | H-Lab Life | “Multi-Use Spray” products |  | https://hlablife.com |
| 8/10/20 | Greater Peoria Physical Medicine & Rehabilitation, S.C. d.b.a. Joseph Health Group | Basic Immune Support Kit, Enhanced Immune Support Kit, and Ultra Immune Support Kit | Dr. Daniel T. Joseph | http://josephhealthgroup.com |
| 8/11/20 | Oxford Medical Instruments USA, Inc. | Salt inhaler products, including "OMI Salt Therapy Pipes" | Istvan Magyar,CEO | https://www.oxfordmedicals.com |
| 8/14/20 | SilveryGuy | Colloidal Silver |  | www.silveryguy.com |
| 8/17/20 | Predictive Biotech | CoreCyteTM | Mr. Eric K. Olson | www.predictivebiotech.com and https://predtechgroup.com |
| 8/17/20 | PA Green Wellness LLC dba A Predictive Biotech Certified Facility | CoreCyte™ |  | www.pagreenwellness.com |
| 8/18/20 | Pomegranate Consulting, LLC, Pomegranate Consulting, Ltd. dba Glorious One-Pot Meals | “COVID-19 test package” | Elizabeth Gail Yarnell | www.elizabethyarnell.com |
| 8/19/20 | Durazo Medical Biomagnetism | “Biomagnetism Magnetic Therapy DIY Kits” | Moses Durazo | www.savememagnets.com |
| 8/19/20 | Living Senior, LLC | CBD products | Justin Hartfield | www.blueribbonhemp.com and www.cbdseniors.com |
| 8/27/20 | Lattice Biologics, Ltd. | Amniotic fluid product (sometimes referred to as AmnioBoost) | Guy Cook,Chief Executive Officer | www.latticebiologics.com, |
| 9/1/20 | 1 Party At A Time | BetterFly, a niacin containing product | Dmitry Kats | https://reternity.org |
| 9/9/20 | Pharmacy Plus, Inc. dba Vital Care Compounder | “COVID PACK” and “COVID ‘POSITIVE’ PACK” | Ron Edwards | www.vitalcarecompounder.com |
| 9/11/20 | KetoKerri LLC | “KK Black Seed Oil,” “KK Breakthrough Vitamin D with Chondroitin & Oleic," “Stonebreaker,” “KK EDTA with Selenium and Minerals,” “Zeolite,” “Ultra Liquid Zeolite,” and “DR. FITT FIRE FIGHTERS” | Kerri Rivera | www.kerririvera.com and www.ketokerri.com |
| 9/22/20 | Nephron Pharmaceuticals Corporation | ANDA 78202 Budesonide Inhalation Suspension, for inhalation suspension |  | http://www.nephronpharm.com/ |
| 9/29/20 | Tonic Therapeutic Herb Shop & Elixir Bar | Herbal products |  | https://www.tonicherbshop.com |
| 10/5/20 | Physician 360, Inc. | Physician 360 COVID-19 Rapid Test | Dr. Rob Lapporte,Co-Founder and CMO | https://physician360.co/ |
| 10/7/20 | Prairie Dawn Herbs | Herbal products | Dawn Brooks | https://www.prairiedawnherbs.com |
| 10/7/20 | Griffo Botanicals | Herbal tincture products | Frank Griffo | https://griffobotanicals.com |
| 10/15/20 | LVWellness & Aesthetics | “COVID-19 ANTIBODY TEST KIT," ViraFend Multi-Virus Defense product | Justin Hogan | www.lvwell.com |
| 10/16/20 | For Our Vets LLC dba Patriot Supreme | CBD products | Justin Elenburg | https://patriotsupreme.com |
| 10/23/20 | Beepothecary LLC | Bee products, including “Elderberry, Honey & Propolis Syrup,” “BEEbread,” and “BEEHive Delight,” | Laurie Dotson/Peter Dotson | www.beepothecary.us and www.beepothecary.square.site |
| 10/23/20 | Peterson Research Laboratories LLC | "Simple Silver” | Robert L. Peterson | www.covercology.com |
| 10/23/20 | Predator Nutrition | Elixir (labeled to contain oxymatrine, berberine, and ecklonia cava), Salidroside, Unbreakable (labeled to contain BPC-157), Vitamin C + Bioflavonoids & Rosehip, Vitamin D3, and Ashwagandha | Rajinder Singh Johal | www.predatornutrition.com |
| 10/29/20 | Trask and Roth Inc. dba Trask & Roth Research and Manufacturing | “Lateral Flow Testing Device” | Kodi Bethay-Roth,President | www.traskandroth.com |
| 10/30/20 | Spartan Enterprises Inc. dba Watershed Wellness Center | “Dissolve BioActive Silicate,” and products labeled to contain silver | Robert McCauley | https://watershed.net |
| 11/2/20 | NovaBay Pharmaceuticals, Inc. | ANTIMICROBIAL LID & LASH SOLUTION, PURE HYPOCHLOROUS ACID, 0.01%, 20 m | Justin Hall,Chief Executive Officer | https://novabay.com, https://avenova.com and https://saniteyesmore.com |
| 11/10/20 | Sage Woman Herbs, Ltd. dba Sage Consulting & Apothecary | X-tra Strength CV Bundle, Golden Mushroom Blend, Golden Mushroom Blend Tincture, Moducare Chewables, Vitamin C with Bioflavonoids,  Protocol for Life Balance K2 MK-7 & D3, Sovereign Silver Bio-Active Silver Hydrosol, Super Elderberry Syrup, Respiratory Tonic, Cistus Incanus Tea, Manuka Honey, Respiratory/Head Lungs, Shuang Huang Lian, Winter Rescue, and Winter Rescue Tincture | Valerie Blankenship | https://www.sagewomanherbs.com |
| 11/17/20 | Innovative Medicine LLC | “Nadovim” | Caspar Szulc | https://innovativemedicine.com/ and https://nadovim.com/ |
| 11/17/20 | ChromaDex | “Tru Niagen products” | Frank Jaksch,Co-Founder | www.chromadex.com and www.truniagen.com |
| 11/18/20 | Pro Breath MD, LLC dba Dentist Select and OraCare | “OraCare Health Rinse” and “OraCare Operatory Pre-Rinsing Set” |  | https://www.oracareproducts.com/ and https://www.dentistselect.net |
| 11/18/20 | Vibrant Health Care, Inc. | Umbilical cord derived cellular product | Edgar Suter, MD | www.vibranthealthcare.org |
| 11/18/20 | Red Moon Herbs | American Ginseng Elixir, Ashwagandha, Astragalus, Breathe Clear, Chickweed, Dandelion, Echinacea, Elderberry, Elderberry Elixir, Elecampane, Garlic Elixir, Ginkgo, Hyssop, Kudzu, Lobelia, Lung Support, Mushroom Elixir, Turmeric, Usnea, Violet, Viral Spiral, Wild Cherry Bark Syrup, and Yarrow | Jeanette Dunn Acevedo | https://redmoonherbs.com/ |
| 11/23/20 | Industry Lab Diagnostic Partners | “COVID-19 Testing Kit” | Lance Benedict,President/CEO | www.ildp.com |
| 11/30/20 | Avazo-Healthcare, LLC | COVID-19 test kit products and CBD products |  | https://www.covidtests.shop and www.cbdmarketweb.com |
| 12/2/20 | Rat's Army | “VIRUS BIOSHIELD" |  | https://rats.army/ |
| 12/2/20 | Health & Wellness Center International One, L.L.C. dba Hotze Vitamins | “Dr. Hotze’s Immune Pak with Vitamins A, B, C, D, Zinc and Probiotics," “Dr. Hotze’s Kids Immune Pak,” and “Dr. Hotze’s Teen Immune Pak” | Dr. Steven F. Hotze, MD | www.hotzehwc.com and www.hotzevitamins.com |
| 12/2/20 | Heavenly Natural Products | C60 (Carbon 60) and colloidal silver products, including “AVOCADO C60 ANTI-VIRAL COMBO – VIRUS PREVENTION” |  | https://heavenlynaturalproducts.com |
| 12/7/20 | Paradigm RE LLC | "Thymosin Alpha 1” |  | https://paradigmpeptides.com/ |
| 12/10/20 | iThrive.health | Vitamins, quercetin, zinc and omega-3 products | Alirio Zavarce | https://ithrive.health/ |
| 12/10/20 | Indigenous Products | Mineral products | Steve Crear | https://www.facebook.com/Indigenous-Products-106070747498475/ |
| 12/18/20 | Rowpar Pharmeceuticals | CloSYS Oral Spray, CloSYS Non-Irritating RINSE UN-flavored for ULTRA SENSITIVE Mouths, and CloSYS Non-Irritating RINSE Mildly Flavored for SENSITIVE Mouths products | James Ratcliff,CEO | www.closys.com |
| 12/21/20 | Riverstone LLC | “Flu Immune Drops,” “L-Lysine,” “Lysine Extra,” and “Monolaurin” | Chris LaVoie | https://www.doctorschoice.org |
| 12/21/20 | Sparrow Health & Performance LLC | Organic Liposomal Vitamin C, Nanoemulsified D3K2, and Immune Support Package (which includes Organic Liposomal Vitamin C, Nanoemulsified D3K2, and Virus Be Gone products, with Smart Silver as an optional add-on) |  | https://www.sparrowclinic.com |
| 1/4/21 | Coco's Holistic Specialties & Apothecary | 4-Thieves Florida Tea Concentrate and 4-Thieves Florida Tea Powder | Janine Havins | https://cocosholisticspecialties.org/ |
| 1/12/21 | AusarHerbs | “Corona Destroyer Tea” |  | https://ausarherbs.com/ |
| 1/12/21 | Allimax Us | Allicin capsules, creams, gels, sprays, and liquid products | Bobbi Walton | www.allimax.us |
| 1/25/21 | EnMed MicroAnalytics, Inc. | “COVID-19 BLOOD SPOT COLLECTION PACKET” and the “SARS-CoV-2 SALIVA RT-PCR TEST COLLECTION PACKET” | Edwin Naylor | https://enmedmicroanalytics.com |
| 1/25/21 | PYRLess Group, LLC dba Dr. Fitt | Buffered C Powder, Get Well 2 Day Vit A, Get Well 2 Day Vit D3, Robynzyme, and OMG Cell Protek | Christy McCord | https://drfitt.com and https://drfittinfo.com |
| 2/16/21 | Dr. Paul's Lab | “COVID-Aid Tincture” | Paul Dettloff, DVM | http://drpaulslab.net |
| 2/18/21 | SafaLab, Inc. | The Prevention 12 Pak (also referred to as the Coronavirus Prevention 12 Pak), consists of the products SOUL, CORE, Cellular Detox, Turb-O2, Vitamin C (Anti-Oxidant Assist), Vitamin D3 (Light Assist), Glutathione, Selenium, Zinc, Vitamin A, Melatonin, and Iod Assist (Iodine) | Dr. Bradford Weeks | www.safalab.com and www.weeksmd.com |
| 2/18/21 | B4B Corp. | Earth Tea Extra Strength |  | https://www.b4bcorp.com |
| 2/18/21 | Mercola.com, LLC | “Liposomal Vitamin C,” “Liposomal Vitamin D3,” and “Quercetin and Pterostilbene Advanced” | Dr. Joseph M. Mercola, DO | https://www.mercola.com and https://www.mercolamarket.com |
| 2/26/21 | Orvic dba Webstore-USA | “COVID-19 BLOOD SPOT COLLECTION PACKET” and the “SARS-CoV-2 SALIVA RT-PCR TEST COLLECTION PACKET” |  | https://facemaskandppestore.com |
| 3/1/21 | Ageless Global, LLC | “Immunoral,” “Immune Plus,” “MD Immune Support Spray,” and “MD CVK-365 Mouth Spray” |  | www.agelessgloballab.com, |
| 3/4/21 | Opgal Optronic Industries Ltd. | “Therm-App MD” and “Therm-App MD Pro” | Eran Bluestein | https://www.opgal.com |
| 3/4/21 | Thermavis | “Multi-Person Thermal Screening Camera” | Mark Norman Nightingale | https://thermavis.com/ |
| 3/4/21 | Thermoteknix System Limited | “FevIR Scan 2” | Richard Sydney Salisbury and Geraldine Anne Salisbury | (http://www.thermoteknix.com/) |
| 3/4/21 | Westminster International Ltd | “WG 520” and “WG 620” | Roger William Worrall | https://www.wi-ltd.com/ and https://www.wg-plc.com/ |
| 3/4/21 | Workswell Group s.r.o. | “MEDICAS” | Adam Svestka | (https://workswell-thermal-camera.com/) |
| 3/4/21 | CreativeStar Solutions | “Artemis TI-CS-T11 Thermal Scanner” and “Artemis Gateway” | Steven Gao/Dan Xiao | https://cssinco.com/ |
| 3/4/21 | Omega Engineering Inc. | “TI-120GTS” |  | https://www.omega.com/ |
| 3/4/21 | Dubak Electrical Group | “DuThermX” | Vlastimir Dubak | https://www.duthermx.com/ and https://www.dubakelectrical.com/ |
| 3/4/21 | Shenzhen Sunell Technology Corporation | Bi-Spectrum Fever Screening Network Camera (SN-T5P-F), Temperature Screening Thermographic Network Bullet Camera (SN-F22-B), AI Fever Screening Network Camera With Integrated Blackbody (SN-T5H-P-F), and Fever Screening Body Temperature Measurement Network Camera Accurate ±0.3_ (SN-T5/F) | Ann Wu | https://www.sunellsecurity.com/ |
| 3/4/21 | Omnisense Systems Pte Ltd | Sentry MK4 |  | https://www.omnisense-systems.com/ |
| 3/5/21 | CAMA Wellness Center/IodoRios Company, LLC | Hand wipe product | Elise and Max Rivers | www.cp.camacenter.com |
| 3/12/21 | Ravenscroft Apothecary, Inc. DBA Ravenscroft Escentials | “AIR PURIFY AROMAMIST,” “HEALER’s CHAI AROMATEA & BREATH DROPS,” “ELEVATION OF MIND AROMAMIST,” and “ROSE FREQUENCY TONGUE TINCTURE” | Ginger Ravenscroft | www.ravenscroftescentials.com |
| 3/18/21 | Block Scientific | “QUIKPACII COVID-19 ANTIBODY TEST KIT” and the “Long Island Biotech COVID-19 Antigen Rapid Test (Colloidal Gold)” | Jeremy Linder | www.blockscientificstore.com |
| 3/24/21 | Tresmonet Technologies, Inc., TM Testing, Inc. dba TM Technologies, Inc./TM Labs | “COVID-19 IgM/IgG Rapid Test” | Dennis Dannel,Founder and CEO | www.tmtestkits.com |
| 3/25/21 | Ikcon Investments, Inc dba Ikcon Medical | “Akcutest COVID-19 Antibody Test” and the “Lumigenik COVID-19 Antigen Rapid Test” | Ricardo Emmanuel Weichsel Upton,President | www.ikconmedical.com and www.akcutestshop.com |
| 4/1/21 | Natural Adventure, LLC | Purity Sanitizer with 70% Alcohol and Purity Essential Oil Blend products | Jennifer Orejobi | www.mynaturaladventure.com |
| 4/6/21 | Sethi Laboratories | “Rapid COVID-19 Antibody Self Test Kit” and a “Rapid COVID-19 Antigen Self Test Kit” | Mr. Sameer Sethi | https://sethilaboratories.com |
| 4/7/21 | Allure Imports | Silver Soul Immune Support, Silver Soul Body Spray and Vitality C60 | Cory Chiarello | www.facedoctor.ca, www.vitalityc60.com, and www.allureimports.com |
| 4/7/21 | About Mineral | Puriton topical skin products | Hyun Eun Lee | https://aboutmineral.com |
| 4/13/21 | Trinity Natural Health & Pain Management, Inc. | “COVID-19 Formula” |  | https://trinity-naturalhealth.com/,https://www.facebook.com/TrinityNaturalSupplements, |
| 4/13/21 | Anytime COVID Test LLC | “Covid-19 Test Kit” | Steve Utley,CEO | https://www.anytimecovidtest.com and http://www.mycovidtester.com, http://www.gotcovidtests.com, http://www.covidtestyou.com, and http://www.buymycovidtest.com |
| 5/4/21 | Baltimore Beauty Security Square Mall | GenBody COVID-19 Ag test and the GenBody COVID-19 IgM/IgG test | Yoo Kim |  |
| 5/6/21 | Disinfect & Shield | DISINFECT™ & SHIELD HAND SANITIZER | Despo Caldwell,CEO | https://www.disinfectandshield.com/ |
| 5/6/21 | Covalon Technologies Inc. | non-alcohol based hand sanitizer products | Brian Pedlar,CEO | https://www.covaguard.com/ |
| 5/19/21 | BGP, LLC | “BIOCENCE WS Multi-use Selective Antibacterial / Antiviral Human OTC Drug” |  | www.biocence.com |
| 5/24/21 | Beauty & Spa Concepts, DBA Beenefits | Brazilian Propolis Extract |  | www.beenefits.com |
| 5/24/21 | Everything Health LLC | CoronaBox | Tamika Moseley | https://ssnaturalhealing.com |
| 5/27/21 | OCLO LLC/OCLO Nanotechnology Science | Various chlorine dioxide products, including "OCLO 3000" | Ricardo Garcia | https://dioxidodecloro.us |
| 6/10/21 | Innova Medical Group, Inc. | SARS-CoV-2 Antigen Rapid Qualitative Test | Daniel J. Elliot,Chief Executive Officer |  |
| 6/22/21 | Pacific Center of Health/Pacific Center of Health & Acupuncture | Immune Builder, High Risk Environment Pack- NO SYMPTOMS, Anti-Viral Herbal Formula, Anti-Viral Herbal Pack ‚Äì ACUTE ILLINESS, Kitchen Sink, Lung Formula Dry Cough, Lung Formula Wet Cough, Colloidal Silver and Post COVID-19 Recovery products | Richard Gold | www.pacificcenterofhealth.com |

**S2. Daily counts of key phrases**

Please see daily_counts.xlsx

**S3. Additional performance details**

Additional products that were detectable if the initial bias was not added:

*Colloidal silver* (FDA letter date: 03/06/2020)

*Essential oil* (FDA letter date: 03/06/2020)

*Super C* (FDA letter date: 04/21/2020)
