## Supplementary figures and images for "Early detection of fraudulent COVID-19 products from Twitter chatter"

### Figure 1-highres.jpg

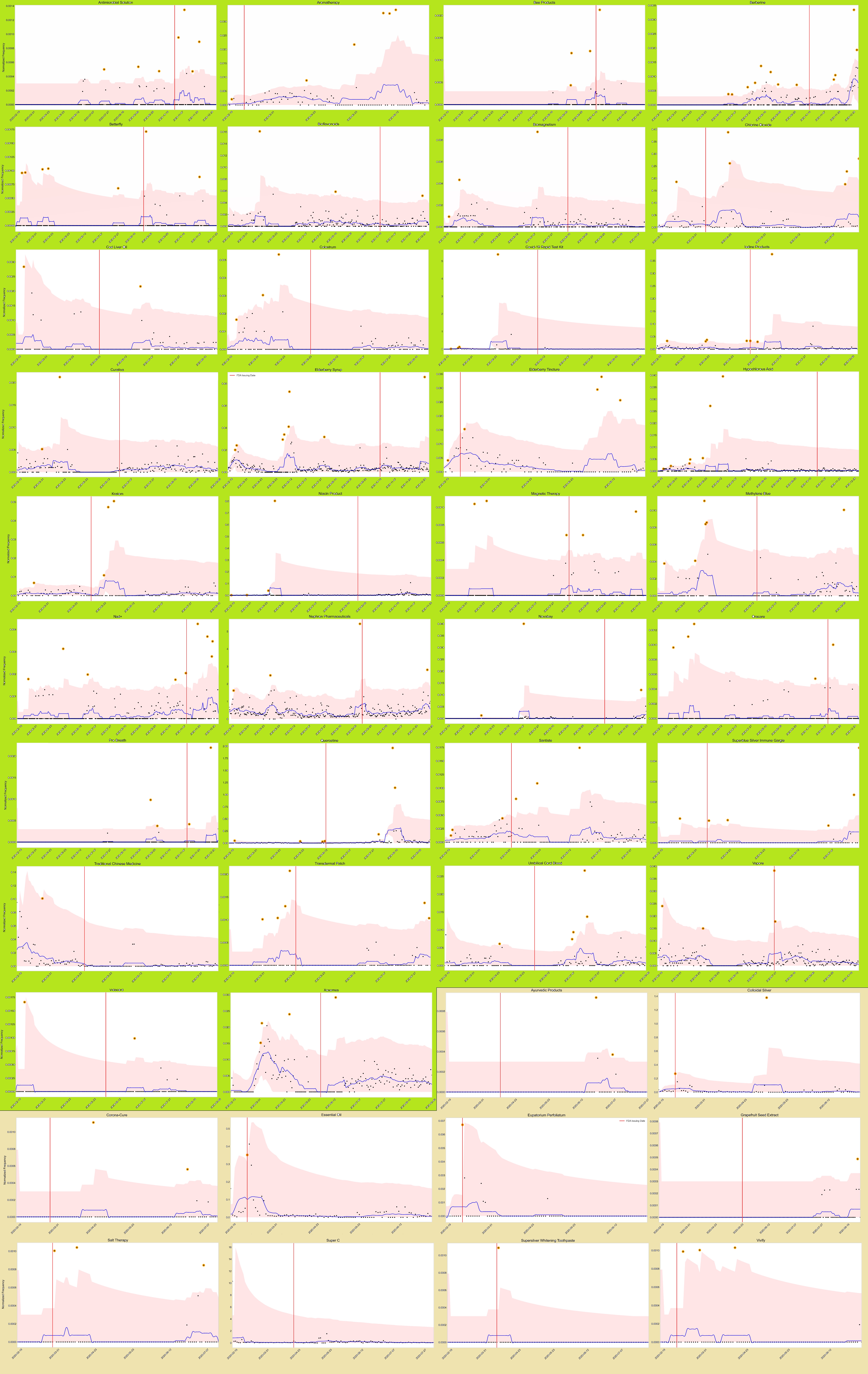
